# Enhancing Tele-Simulation during OSCE: Incorporating Feedback from Standardized Patients

**DOI:** 10.1101/2021.10.05.21264554

**Authors:** Meghana Sudhir

## Abstract

**Background:** As COVID-19 impacted universities across the globe for over one year, the organization of OSCEs also demanded change. The traditional OSCEs had to transition to Tele simulation OSCEs. As the pandemic continues, there is possibility that this approach may continue for some time, it is imminent to capture feedback from the simulated patients so that we can explore ways to improve in future.

**Objectives:** To gather feedback from standardized patients doing Tele Simulation in OSCEs, To develop plans to improve Tele Simulation involving Standardized Patients based on the feedback gathered.

**Method:** Data were collected immediately after Tele simulation OSCE during debriefing from 18 Simulated Patients. This was followed up with an open-ended questionnaire and gathered additional information.

**Results:** The feedback from SPs included their feelings when they were introduced to Tele simulation OSCE, Tele simulation versus Face to Face OSCE and areas of improvement.

**Conclusion:** Tele simulation OSCE was a quick adaptation during COVID-19 pandemic. But the lessons learned, and the feedback received are pearls for future innovative approaches to assessments. It is encouraged to collect verbal and written feedback for the improvement of Tele simulation.

## Introduction

Simulated Patients (SP) have been contributing to health professions education for decades. Assessment is an important area where the simulated patients take the role of standardized patients [1]. They play their role consistently to replicate the same experience with all examinees. Over the years these were face to face encounters in a simulated setting conducted as Objective Structured Clinical Examination (OSCEs).

As COVID-19 impacted universities across the globe for over one year, the organization of OSCEs also demanded change [2]. The traditional OSCEs had to transition to Tele simulation OSCEs. Tele simulation can be defined as a process in which telecommunication and simulation resources are used to provide education, training, and/or assessment to learners at an off-site location [3].

Tele simulation also needed to readjust to the demands of the situation [4]. At our university, to maintain safety of everyone involved, the number of people in each station was reduced to two. Only examiner and candidate were inside the station and SPs were in separate rooms doing Tele Simulation. Candidate and SPs met online through laptop rather than face to face interaction. The examiner made the scoring based on this interaction.

As the pandemic continues, there is possibility that this approach may continue for some time, it is imminent to capture feedback from the standardized patients so that we can explore ways to improve in future.

### Objectives

- To gather feedback from standardized patients doing Tele Simulation in OSCEs
- To develop plans to improve Tele Simulation involving Standardized Patients based on the feedback gathered.

## Method

Feedback from the SPs were gathered after the Foundations of Clinical Medicine OSCE. As with every simulation activity, debriefing is utmost important; the same paved to be beneficial in gathering feedback from the SPs after Tele OSCE [5].

18 SPs took part in Foundations of Clinical Medicine OSCE. At the end of OSCE, a structured debriefing was conducted. Gibbs reflection model was used for debriefing. [6] Figure 1. The information gathered from the debriefing gave immediate feedback after the OSCE.

**Figure 1:**
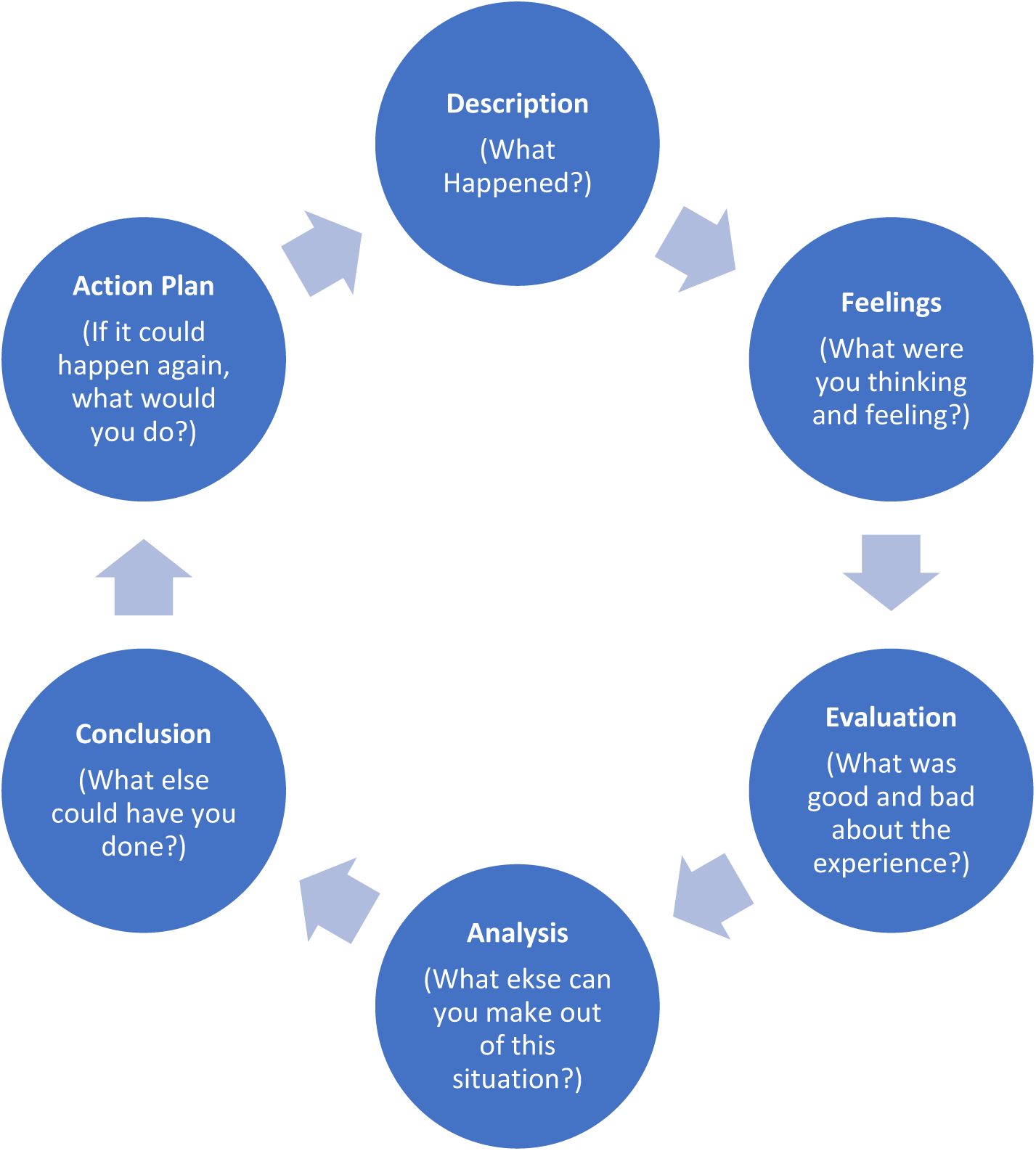
Gibbs Reflection Model

The Standardized patients also received an online questionnaire. Any further information which they missed in the debriefing was captured using that questionnaire.

## Summary of Results

### Being an SP in Tele simulation OSCE –Feelings

Some of the feelings shared by SPs are shared in Figure 2

**Figure 2:**
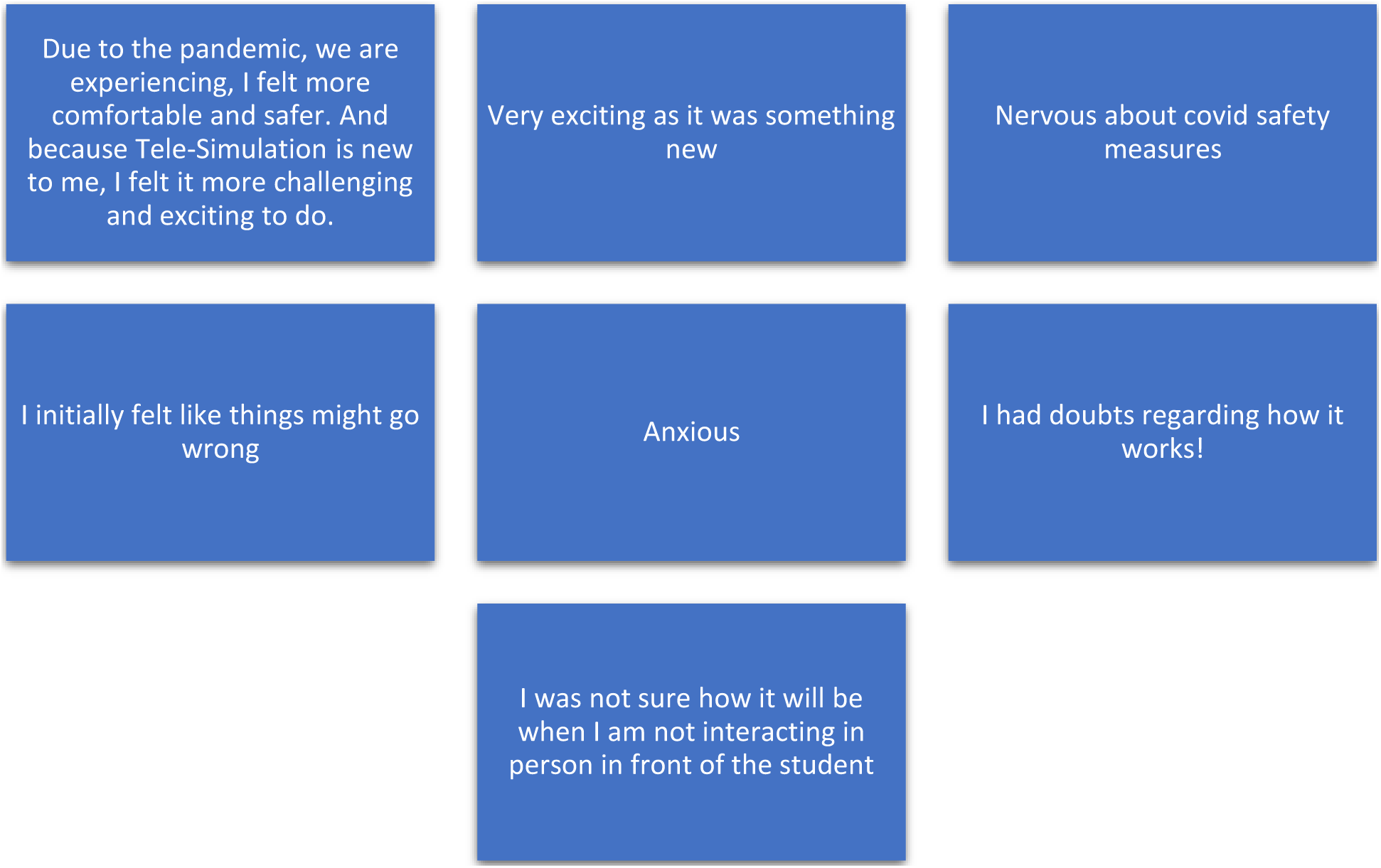
Being an SP in Tele simulation OSCE –Feelings

### Tele Simulation Versus Face-to-Face Simulation in OSCE

Some of the responses from SPs are shared below

- *“In response to Covid-19 outbreak, I found Tele Simulation more helpful and safe over the Face-to-Face Simulation because social distancing was well implemented*.*”*
- *“There was obviously a lack of human connection but considering the circumstances it was a very safe and convenient way of doing things. Online doctor appointments are actually becoming the way of the future, so it is very good to train these future doctors for it*.*”*
- *“Both are the same for me personally. But I felt I could be more expressive with my emotions when face to face*.*”*
- *“Both are important given the current context and future of medicine. OSCE must have 30% tele & 70% face to face simulations”*.
- *“I think both are okay, considering whatever situation is on ground for instance with Covid 19, the best option is the Tele Simulation”*.

### Which type of simulation in OSCE You prefer? Tele Simulation or Face to Face? Why?

SPs had mixed response to the preference for the simulation modality for OSCE.

- *“I found both the ways have their pros and cons but personally I did miss the physicality of the experience so I would have to say face to face”*.
- *“Face to face is preferred* … *technological difficulties can be avoided especially during the exam”*.
- *“Face to face as acting out the scenario would be more easy with direct interaction with the students*.*”*
- *“I, personally don’t mind acting any of the two, but I will prefer face to face”*
- *“Both are fine with me”*

### Plans to improve Tele simulation OSCE from the feedback gathered

This was the first Tele simulation OSCE during the pandemic. The OSCE team worked together to make the experience as realistic and smooth as possible with the available resources. The SPs provided firsthand feedback after the Tele simulation OSCE which helped to improve for future Tele simulation sessions.

Some of the improvement areas SPs shared are as follows

- *“Some laptops provided had cameras at the left bottom corner. It was difficult to keep eye contact with the student as well as to look at the camera at the bottom. Need to have laptops with camera at the top center*.*”*
- *“The students also need to be briefed on how to position themselves for the Tele OSCE and how to maintain eye contact during the interview*.*”*
- *“The training provided with technical requirements for Tele OSCE was helpful. It is important to train SPs with the logistics of Tele simulation along with role play scenario*.*”*
- *“As this Tele simulation OSCE had no scoring from SPs, the organization was fine. But I am not sure how to score a candidate through this platform. This could be an area for improvement for future sessions*.*”*
- *“Body language has restriction when OSCE is done as Telesimulation. So, facial expression, modulation of voice, eye contact etc need more emphasis*.*”*

## Discussion

Tele simulation OSCE was a novel experience for all SPs who took part in this OSCE. It was a new experience for the organizing team also. Feedback being one of the important features of simulation, which is essential to promote effective learning, the experiences shared by SPs contributed to subsequent adaptations in future assessments [7]. Tele simulation provided value and flexibility in which simulation education could continue during the unprecedented times [8].

The difficulty to express nonverbal communication was a challenge with SPs with similar restrictions on the candidate’s nonverbal communication noticed [9]. This is an area where the SPs need to be aware of this restriction. As mentioned by one of the SPs, the communication can be improved by enhancing voice modulation, facial expression, and eye contact. SP educators need to train SPs with emphasis to non verbal cues. The candidates also need orientation and support in developing their verbal and non-verbal communication skills in Tele simulation OSCE [10].

Telemedicine is another area where Tele simulation OSCEs are significant [11]. The feedback from some of the SPs indicated that they are aware of the possibility of them participating in Tele OSCEs in Telemedicine. With COVID-19 pandemic continues, need for further Telemedicine consultations seems empirical.

Majority of SPs shared their difficulty with ‘nose-cam’ issue of laptop where the camera was at the bottom corner rather than at the top center. As the SPs had to look at the camera, they were unable to look at the candidate on screen. This was rectified for subsequent sessions. It is important to see that the laptops provided for Tele Simulation OSCE should have cameras at the top.

Training of SPs for Tele simulation OSCE needs additional emphasis on technology. SPs working with Telemedicine OSCEs are trained according to the need of scenarios at organizations [12]. This was not the case with the SPs at our institution. They were trained on scenarios as well as the IT team was on site for clarifying their queries during training. This helped in the overall implementation of Tele simulation OSCE. A collaborative training plan incorporating IT basics for Tele simulation OSCE is highly recommended.

Some OSCE stations require SPs to score candidates. The scorings are based on the reflection on “how well you feel you were treated”[14]. During the Tele simulation OSCE, even though the SPs were not asked to score the candidates, some of the SPs shared their concern regarding how to score on an online platform. This was taken into consideration and a separate chat option was created between the SP and examiner. The SP could score on the chat window and the examiner entered it on the scoring platform.

SPs felt safe during COVID-19 times doing Tele simulation OSCE. ‘Safe work environment’ - Domain 1 of ASPE Standards of Best Practice was emphasized [14].They were provided with well-lit and well-ventilated individual rooms with access to all safety measures. Safety is the corner stone of simulation.

## Conclusion

Tele simulation allows the benefits of simulation to extend beyond the walls of a simulation center [3]. Tele simulation OSCE was a quick adaptation during uncertain times. With the COVID-19 situation getting better, the OSCEs are back to face to face encounters with adequate safety protocols. But the lessons learned, and the feedback received are pearls for future innovative approaches to assessments. It is encouraged to collect verbal and written feedback for the improvement of Tele simulation [15]. A 360 degree feedback from everyone involved in Tele Simulation OSCE -Examiners, SPs, Candidates, Administrative personnel, IT Team and others will definitely support the organizations in future planning of Tele simulation sessions.

## Data Availability

The data is secured in password protected computer at Mohammed Bin Rashid University of Medicine and Health Sciences

## Ethics Approval

The Institutional Review Board at Mohammed Bin Rashid University of Medicine and Health Sciences approved the study proposal on 25 January 2021. Reference: MBRU IRB-2020-20

## External Funding

None

## Acknowledgements and Disclosures

I acknowledge the contributions of all participants as well as the support of Simulation Center team in the completion of this project. This work was supported in part by the Al Jalila Foundation.

